# Integrating a host transcriptomic biomarker with a large language model for diagnosis of lower respiratory tract infection

**DOI:** 10.1101/2024.08.28.24312732

**Authors:** Hoang Van Phan, Natasha Spottiswoode, Emily C. Lydon, Victoria T. Chu, Adolfo Cuesta, Alexander D. Kazberouk, Natalie L. Richmond, Padmini Deosthale, Carolyn S. Calfee, Charles R. Langelier

**Affiliations:** Department of Medicine, Division of Infectious Diseases, University of California San Francisco; Department of Pediatrics, Division of Infectious Diseases and Global Health, University of California San Francisco; Chan Zuckerberg Biohub San Francisco; Department of Medicine, University of California San Francisco; Department of Medicine, Division of Pulmonary, Critical Care, Allergy and Sleep Medicine, University of California San Francisco

## Abstract

**BACKGROUND:** Lower respiratory tract infections (LRTIs) are a leading cause of mortality worldwide and can be difficult to diagnose in critically ill patients, as non-infectious causes of respiratory failure can present with similar clinical features.

**METHODS:** We developed a LRTI diagnostic method combining the pulmonary transcriptomic biomarker *FABP4* with electronic medical record (EMR) text assessment using the large language model Generative Pre-trained Transformer 4 (GPT-4). We evaluated this approach in a prospective cohort of critically ill adults with acute respiratory failure from whom tracheal aspirate *FABP4* expression was measured by RNA sequencing. Patients with LRTI or non-infectious conditions were identified using retrospective, multi-physician clinical adjudication. We then confirmed our findings by applying this method to an independent validation cohort of 115 adults with acute respiratory failure.

**RESULTS:** In the derivation cohort, a combined classifier incorporating *FABP4* expression and GPT-4– assisted EMR analysis achieved an AUC of 0.93 (±0.08) and an accuracy of 84%, outperforming *FABP4* expression alone (AUC 0.84 ± 0.11) and GPT-4–based analysis alone (AUC 0.83 ± 0.07). By comparison, the primary medical team’s admission diagnosis had an accuracy of 72%. In the validation cohort, the combined classifier yielded an AUC of 0.98 (±0.04) and an accuracy of 96%.

**CONCLUSIONS:** Integrating a host transcriptional biomarker with EMR text analysis using a large language model may offer a promising new approach to improving the diagnosis of LRTIs in critically ill adults.

**Description:** We present the novel use of a host transcriptional biomarker combined with artificial intelligence analysis of electronic medical record data to diagnose lower respiratory tract infections in a derivation cohort of critically ill adults, then the validation of this approach in a second, fully independent, cohort. This approach demonstrated high diagnostic accuracy compared to a gold standard of post-hoc multi-physician adjudication.

## INTRODUCTION

Lower respiratory tract infections (LRTIs) are a leading cause of death worldwide, yet remain challenging to diagnose^1^. This is especially true in the intensive care unit (ICU), where non-infectious acute respiratory illnesses often have similar clinical manifestations. Further complicating accurate diagnosis is the failure to identify a causative pathogen in most clinically recognized cases of LRTI^2^. The resulting diagnostic uncertainty drives the overuse of empiric antibiotics, leading to adverse outcomes ranging from *Clostridioides difficile* infection to the development of antimicrobial resistance^3,4^.

Host transcriptional biomarkers are a promising modality for LRTI diagnosis that overcome several limitations of traditional microbiologic tests^5,6^. By offering a more direct and dynamic measure of the host immune response, they can enable earlier and more accurate identification of infection, and differentiate between bacterial and viral causes of pneumonia, even in cases where pathogen detection is challenging^1,2^. Single gene biomarkers are particularly amenable to clinical translation, as they can be readily incorporated into nucleic acid amplification platforms widely used in healthcare settings.

Pulmonary *FABP4*, for instance, was recently identified as a novel LRTI diagnostic biomarker in critically ill patients with acute respiratory failure, achieving an area under the receiver operating characteristic curve (AUC) of 0.85 ± 0.12 in adults^7^. Despite better performance characteristics than existing clinical protein biomarkers such as C-reactive protein^8^ or procalcitonin^9^, *FABP4*, like most pneumonia diagnostic biomarkers, may not yet provide the accuracy needed to enable confident clinical decisions regarding antimicrobial use in ICU patients with acute respiratory failure.

Given that large language models (LLMs) such as Generative Pre-trained Transformer 4 (GPT-4) have shown promise in a diversity of medical applications^10^, we considered the possibility that they could be leveraged to improve host biomarker-based LRTI diagnosis. LLMs have demonstrated remarkable performance for image interpretation^11^, patient risk stratification^12^, and assisting with clinical reasoning^13-15^, although their utility for diagnosing LRTI or other critical illness syndromes has not been assessed. Here, we address this gap by building a diagnostic classifier combining *FABP4* with GPT-4 analysis of electronic health record (EHR) data. We find that this combination affords remarkably accurate LRTI diagnosis, suggesting a promising new approach to improve the care of critically ill patients.

## METHODS

### COHORTS AND ADJUDICATION OF LRTI STATUS

We studied patients from two prospective observational cohorts of critically ill adults with acute respiratory failure enrolled at the University of California San Francisco (UCSF) Medical Center (**Figure 1, Table 1**). All patients were enrolled within 72 hours of intubation under UCSF Institutional Review Board protocols #10-02701 (derivation cohort^16^, N=202; enrolled 10/2013-01/2019) or #20-30497 and #10-02852 (validation cohort; N=115; enrolled 04/2020-12/2023).

**Figure 1.**
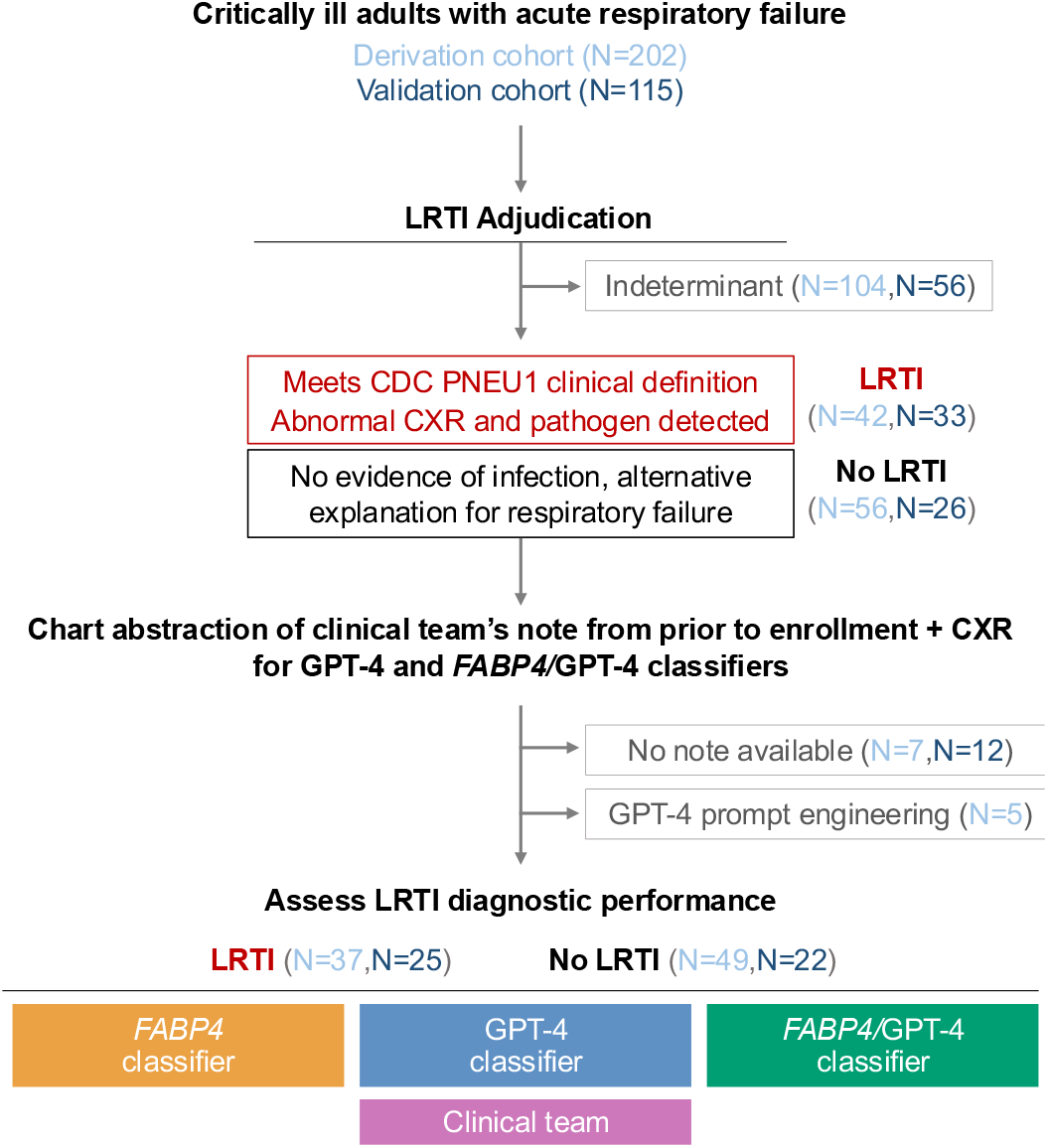
Study flow diagram and overview. Abbreviations: LRTI = lower respiratory tract infection; RNA-seq = RNA sequencing; CXR = chest X ray, FABP4 = gene encoding fatty acid binding protein 4; CDC = U.S. Centers for Disease Control and Prevention; GPT-4 = Generative Pre-trained Transformer 4.

**Table 1.**
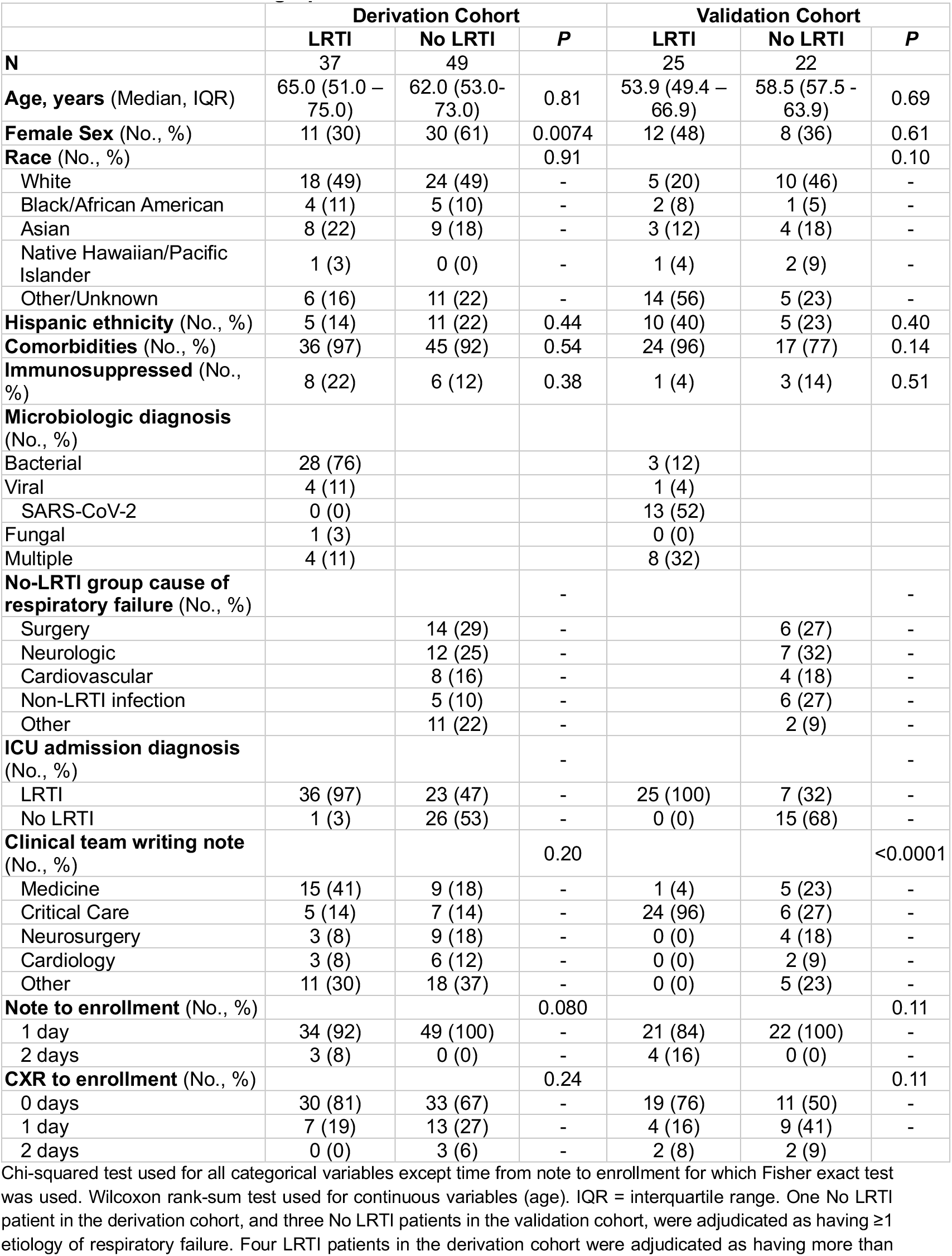
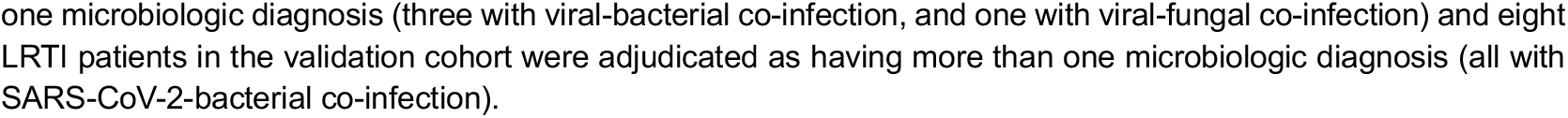
Clinical and demographic features of cohorts.

|  | Derivation Cohort |  |  | Validation Cohort |  |  |
| --- | --- | --- | --- | --- | --- | --- |
|  | LRTI | No LRTI | P | LRTI | No LRTI | P |
| **N** | 37 | 49 |  | 25 | 22 |  |
| **Age, years** (Median, IQR) | 65.0 (51.0 – 75.0) | 62.0 (53.0–73.0) | 0.81 | 53.9 (49.4 – 66.9) | 58.5 (57.5 – 63.9) | 0.69 |
| **Female Sex** (No., %) | 11 (30) | 30 (61) | 0.0074 | 12 (48) | 8 (36) | 0.61 |
| **Race** (No., %) |  |  | 0.91 |  |  | 0.10 |
| White | 18 (49) | 24 (49) | - | 5 (20) | 10 (46) | - |
| Black/African American | 4 (11) | 5 (10) | - | 2 (8) | 1 (5) | - |
| Asian | 8 (22) | 9 (18) | - | 3 (12) | 4 (18) | - |
| Native Hawaiian/Pacific Islander | 1 (3) | 0 (0) | - | 1 (4) | 2 (9) | - |
| Other/Unknown | 6 (16) | 11 (22) | - | 14 (56) | 5 (23) | - |
| **Hispanic ethnicity** (No., %) | 5 (14) | 11 (22) | 0.44 | 10 (40) | 5 (23) | 0.40 |
| **Comorbidities** (No., %) | 36 (97) | 45 (92) | 0.54 | 24 (96) | 17 (77) | 0.14 |
| **Immunosuppressed** (No., %) | 8 (22) | 6 (12) | 0.38 | 1 (4) | 3 (14) | 0.51 |
| **Microbiologic diagnosis** (No., %) |  |  |  |  |  |  |
| Bacterial | 28 (76) |  |  | 3 (12) |  |  |
| Viral | 4 (11) |  |  | 1 (4) |  |  |
| SARS-CoV-2 | 0 (0) |  |  | 13 (52) |  |  |
| Fungal | 1 (3) |  |  | 0 (0) |  |  |
| Multiple | 4 (11) |  |  | 8 (32) |  |  |
| **No-LRTI group cause of respiratory failure** (No., %) |  |  | - |  |  | - |
| Surgery |  | 14 (29) | - |  | 6 (27) | - |
| Neurologic |  | 12 (25) | - |  | 7 (32) | - |
| Cardiovascular |  | 8 (16) | - |  | 4 (18) | - |
| Non-LRTI infection |  | 5 (10) | - |  | 6 (27) | - |
| Other |  | 11 (22) | - |  | 2 (9) | - |
| **ICU admission diagnosis** (No., %) |  |  | - |  |  | - |
| LRTI | 36 (97) | 23 (47) | - | 25 (100) | 7 (32) | - |
| No LRTI | 1 (3) | 26 (53) | - | 0 (0) | 15 (68) | - |
| **Clinical team writing note** (No., %) |  |  | 0.20 |  |  | <0.0001 |
| Medicine | 15 (41) | 9 (18) | - | 1 (4) | 5 (23) | - |
| Critical Care | 5 (14) | 7 (14) | - | 24 (96) | 6 (27) | - |
| Neurosurgery | 3 (8) | 9 (18) | - | 0 (0) | 4 (18) | - |
| Cardiology | 3 (8) | 6 (12) | - | 0 (0) | 2 (9) | - |
| Other | 11 (30) | 18 (37) | - | 0 (0) | 5 (23) | - |
| **Note to enrollment** (No., %) |  |  | 0.080 |  |  | 0.11 |
| 1 day | 34 (92) | 49 (100) | - | 21 (84) | 22 (100) | - |
| 2 days | 3 (8) | 0 (0) | - | 4 (16) | 0 (0) | - |
| **CXR to enrollment** (No., %) |  |  | 0.24 |  |  | 0.11 |
| 0 days | 30 (81) | 33 (67) | - | 19 (76) | 11 (50) | - |
| 1 day | 7 (19) | 13 (27) | - | 4 (16) | 9 (41) | - |
| 2 days | 0 (0) | 3 (6) | - | 2 (8) | 2 (9) | - |

Chi-squared test used for all categorical variables except time from note to enrollment for which Fisher exact test was used. Wilcoxon rank-sum test used for continuous variables (age). IQR = interquartile range. One No LRTI patient in the derivation cohort, and three No LRTI patients in the validation cohort, were adjudicated as having ≥1 etiology of respiratory failure. Four LRTI patients in the derivation cohort were adjudicated as having more than
one microbiologic diagnosis (three with viral-bacterial co-infection, and one with viral-fungal co-infection) and eight LRTI patients in the validation cohort were adjudicated as having more than one microbiologic diagnosis (all with SARS-CoV-2-bacterial co-infection).

Adjudication of LRTI status was performed retrospectively following ICU discharge by two or more physicians using all available information in the EMR, and based on the U.S. Centers for Disease Control and Prevention (CDC) PNEU1 criteria^17^. Patients with a clear alternative reason for their acute respiratory failure besides pulmonary infection, representing the clinically relevant control group, were also identified (No LRTI group). Any adjudication discrepancies were resolved by a third physician, and patients with indeterminate LRTI status were excluded.

### EXTRACTION OF EMR DATA

The primary medical or ICU team’s clinical note from the day prior to study enrollment and the chest X-ray (CXR) read from the day of enrollment were extracted from the EMR. If no note was written on the day prior to enrollment, a note from two days prior was substituted (**Table 1**). If no CXR was performed on the day of enrollment, the next closest CXR read prior to the date of enrollment was used instead. Patients with no clinical notes available prior to study enrollment were excluded (N=7 derivation cohort, N=12 validation cohort). The clinical treatment team’s ICU admission LRTI diagnosis was extrapolated based on administration of antibiotics for empiric treatment of LRTI within one day of study enrollment, excluding antibiotics for established non-pulmonary infections.

### RNA SEQUENCING

RNA was extracted from tracheal aspirate collected on the day of enrollment and underwent rRNA depletion followed by library preparation using the NEBnext Ultra 2 kit on a Beckman-Coulter Echo liquid handling instrument, as previously described^7^. Finished libraries underwent paired-end sequencing on an Illumina NovaSeq.

### *FABP4* DIAGNOSTIC CLASSIFIER

*FABP4* expression was normalized using the varianceStabilizingTransformation function from DESeq2 package (v1.42.1)^18^, and used to train a logistic regression classifier. In each iteration of k-fold cross-validation, both training and test sets were filtered to retain only genes with at least 10 counts across 20% of the samples in the training set. The test fold’s *FABP4* expression level was normalized using varianceStabilizingTransformation and the dispersions of the training folds, and input to the trained logistic regression classifier to assign LRTI or No LRTI status for each patient in the test fold. The performance and receiver operating characteristic (ROC) curve for each of the five folds was evaluated using the package pROC v1.18.5^19^. The mean AUC and standard deviation were calculated from the average AUC derived from each test fold. The sensitivity and specificity at the Youden’s index were extracted for each test fold separately using the function coords from the pROC package, and the average and standard deviation was calculated across the cross-validation folds.

### GPT-4 INPUT, SCORING, AND PROMPT ENGINEERING

We used the GPT-4 turbo model with 128k context length and a temperature setting of 0.2, implemented in Versa, a University of California San Francisco (UCSF) Health Insurance Portability and Accountability Act-compliant model. For each patient, compiled clinical notes and CXR reads were input into the GPT-4 chat interface. Prompt engineering was initially carried out by iterative testing on clinical notes and CXR reads from five randomly selected patients in the derivation cohort, who were excluded from further analyses. We employed a chain-of-thought prompt strategy^20^ that involved asking GPT-4 to analyze the note and CXR step-by-step. The validation cohort included patients enrolled during the height of the COVID-19 pandemic and thus we redacted the terms “SARS-CoV-2” or “COVID-19” from their notes to avoid biasing the GPT-4 analysis. In our final version of the prompt (**Supplement, Appendix 1**), we asked GPT-4 to choose either LRTI or no LRTI, as exemplified in (**Supplement, Appendix 2**). For each patient, GPT-4 was asked to diagnose LRTI in three separate chat sessions. A per-patient GPT-4 score was calculated based on the total number of LRTI-positive diagnoses made by GPT-4.

### INTEGRATED CLASSIFIER

The integrated classifier’s performance was tested using 5-fold cross-validation in the derivation cohort. Because of the smaller sample size, 3-fold cross-validation was used in the validation cohort. For each test fold, a logistic regression classifier was trained on the remaining training folds using both normalized *FABP4* expression and the GPT-4 score. The performance and ROC curve for each fold was evaluated as described above. The sensitivity, specificity and accuracy were calculated based on whether the out-of-fold predicted probability of LRTI is greater than or equal to 50%.

### COMPARING GPT-4 TO PHYSICIANS PROVIDED THE SAME DATA

We compared LRTI diagnosis by GPT-4 against LRTI diagnosis made by three physicians trained in internal medicine (ADK) or additionally subspecializing in infectious diseases (AC, NLR). The physicians were provided identical information and prompt as GPT-4, and they were asked to assign each patient as either LRTI or No LRTI. The comparison physician group score (0 to 3) was calculated based on the total number of LRTI-positive diagnoses made by the comparison physicians.

### DATA AND CODE AVAILABILITY

The gene count data are available at https://github.com/infectiousdisease-langelier-lab/LRTI_FABP4_GPT4_classifier. The code and required source data are available at https://github.com/infectiousdisease-langelier-lab/LRTI_FABP4_GPT4_classifier.

## RESULTS

We evaluated the performance of four different diagnostic approaches (*FABP4*, GPT-4, integrated *FABP4/*GPT-4 classifier, and admission diagnosis by the primary medical team) against a gold-standard of retrospective LRTI adjudication performed by two or more physicians. In the derivation cohort, this adjudication process identified 42 patients with LRTI and 56 with no evidence of infection and a clear alternative explanation for respiratory failure (No LRTI group) (**Figure 1**). In the validation cohort, 33 LRTI and 26 No LRTI patients were identified.

We provided GPT-4 with practical clinical summary information from the EMR that would be available to a treating physician on the day of ICU care: a CXR radiology report from the day of enrollment, and a note written by the medical team from the day prior. In the derivation cohort, notes and radiology reports from five patients were utilized for GPT-4 prompt engineering and optimization (Methods) and seven lacked a clinical note from the day prior to study enrollment, leaving a total of 37 LRTI and 49 No-LRTI cases available for analysis.

We first compared the accuracy of the primary medical team’s ICU admission diagnosis against the gold-standard retrospective LRTI adjudication. The medical team correctly identified 36/37 (97%) of true LRTI cases but incorrectly called LRTI in 23/49 (47%) of patients in the No LRTI group, equating to an accuracy of 72% (**Figure 2A, Table 1**). We next assessed the diagnostic performance of *FABP4* and found that it achieved an AUC of 0.84 ± 0.11 (mean ± standard deviation) by five-fold cross validation (**Figure 2B**). We then assessed the performance of GPT-4 to diagnose LRTI, with three independent diagnoses per patient. A logistic regression classifier based on the GPT-4 score achieved an AUC of 0.83 ± 0.07 (**Figure 2B**).

**Figure 2.**
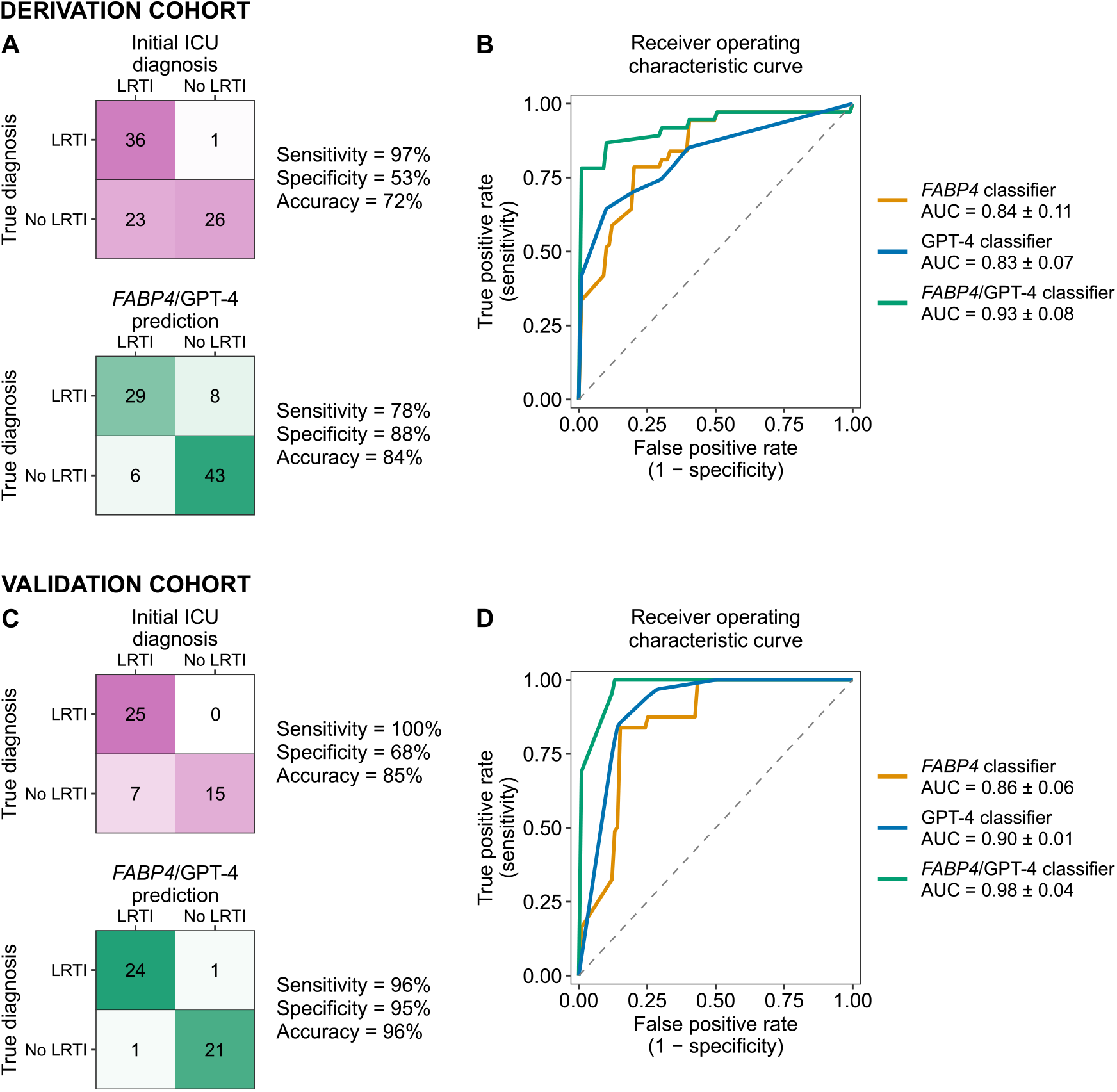
Performance of *FABP4*, GPT-4 and integrated LRTI diagnostic classifiers in the derivation and validation cohorts. **A)** Confusion matrices for initial ICU diagnosis and the integrated *FABP4/*GPT-4 classifier in the derivation cohort. **B)** Receiver operating characteristic curves from GPT-4 classifier, *FABP4* classifier, and integrated *FABP4/*GPT-4 classifier in the derivation cohort. **C)** Confusion matrices for initial ICU diagnosis and the integrated *FABP4/*GPT-4 classifier in the validation cohort. **D)** Receiver operating characteristic curves from GPT-4 classifier, *FABP4* classifier, and integrated *FABP4/*GPT-4 classifier in the validation cohort. In panels A and C, the classifiers output an LRTI diagnosis if the patients had a predicted out-of-fold LRTI probability of 50% or higher. In panels B and D, the area under the curves (AUCs) are presented as mean ± standard deviation.

We then combined *FABP4* and GPT-4 in a single logistic regression model and found that this integrated classifier achieved an AUC of 0.93 ± 0.08 (**Figure 2B**), outperforming both *FABP4* (P = 0.002, one-sided paired t-test) and GPT-4 alone (P = 0.008, one-sided paired t-test). Considering an out-of-fold probability of 50% as LRTI-positive, the integrated *FABP4/*GPT-4 classifier had a sensitivity of 78%, specificity of 88%, and accuracy of 84% (**Figure 2A**). Assessment of the integrated classifier’s performance at the Youden’s index within each cross-validation fold demonstrated an average sensitivity of 86%, specificity of 98%, and accuracy of 93%.

Next, we assessed the validation cohort (**Figure 1**), in which the primary medical team correctly identified 25/25 (100%) of LRTI cases but unnecessarily treated for LRTI in 7/22 (32%) of patients in the No LRTI group, equating to an accuracy of 85% **(Figure 2C)**. The integrated *FABP4/*GPT-4 classifier achieved a sensitivity of 96%, specificity of 95%, and accuracy of 96%, again outperforming either *FABP4* (accuracy 79%) or GPT-4 alone (accuracy 79%). In the validation cohort, the integrated classifier achieved an AUC of 0.98 ± 0.04 using 3-fold cross-validation, as compared to *FABP4* (0.86 ± 0.06) or GPT-4 (0.90 ± 0.01) alone (P = 0.08 and P = 0.02, respectively, one-sided paired t-test) (**Figure 2D)**.

To gain insight into how GPT-4 returns diagnoses based on limited information, we compared the LLM against the decision making of three comparison physicians provided identical input. From the same limited EMR data and prompt provided to GPT-4, we asked the comparison physicians to assign a diagnosis of LRTI or no evidence of LRTI for each patient in the derivation cohort. Considering a threshold of at least one LRTI diagnosis per patient across the three physicians as LRTI-positive, we found a sensitivity of 78%, specificity of 88%, and accuracy of 84% **(Figure 3A)**.

**Figure 3.**
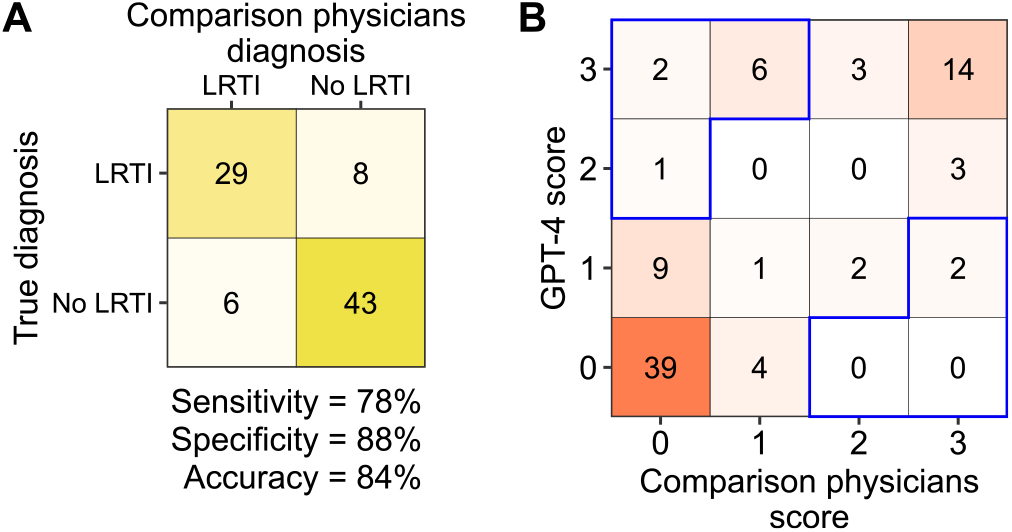
Comparison of GPT-4 performance to physicians provided the same EMR data from the LRTI derivation cohort. **A)** Confusion matrix of diagnosis by three GPT-4 comparison physicians who received the same prompt and data as GPT-4. **B)** Comparison of GPT-4 LRTI scores as compared to physicians. In Panel B, X-axis depicts the number of times GPT-4 diagnosed LRTI out of 3, Y-axis shows the number of times the physicians called LRTI out of 3. Blue boxes indicate instances in which GPT-4 diagnoses were most discordant with comparison physicians (the scores differ by 2 or more).

Considering a threshold of at least one LRTI diagnosis per patient across the three physicians as LRTI-positive, we found a sensitivity of 78%, specificity of 88%, and accuracy of 84% **(Figure 3A)**. Finally, we sought to identify potential biases in GPT-4 diagnoses by comparing GPT-4 results to those of the comparison physicians (**Figure 3B)**, focusing on cases with two or more discordant LRTI diagnoses. Of the nine patients more frequently diagnosed with LRTI by GPT-4 versus the comparison physicians (**Figure 3B**), six had clinical notes with no mention of LRTI, but explicit concern for LRTI in the CXR report. This suggested that GPT-4 may have placed more weight on CXR reads relative to physicians. Of the two patients disproportionately diagnosed with LRTI by comparison physicians versus GPT-4 (**Figure 3B**), one had a final diagnosis of e-cigarette/vaping associated lung injury, and the other had LRTI attributed to rhinovirus.

## DISCUSSION

Our findings demonstrate that the combination of a host transcriptomic biomarker with AI assessment of EMR text data can improve LRTI diagnosis in critically ill patients. We found that an integrated *FABP4/*GPT-4 classifier achieved higher LRTI diagnostic accuracy than *FABP4* alone, GPT-4 alone, or the treating medical team. In our study population, we found that the initial treating physicians unnecessarily prescribed antibiotics in a third to half of patients ultimately found to have non-infectious causes of acute respiratory failure. Had our integrated classifier results been theoretically available at time of ICU admission, we estimate that inappropriate antibiotic use could have been prevented in >90% of No LRTI patients who were unnecessarily treated. Acute respiratory illness is a leading reason for inappropriate antibiotic use^21^, and our results suggest a potential role for biomarker/AI classifiers in antimicrobial stewardship, a major goal of the U.S. CDC^22^ and the World Health Organization^23^.

Previous studies have found that GPT-4 is influenced by the precise language used in a prompt, leading to a need for prompt engineering^14^. By iterating our prompt on a subset of patients, and through direct comparison to physicians provided identical EMR data, we identified possible blind spots of GPT-4 and gained insights that may help guide future optimization of LLMs for infectious disease diagnosis.

A primary strength of this study is the novel combination of a host transcriptional biomarker with AI interpretation of EMR text data to advance infectious disease diagnosis. We address one of the most common and challenging diagnostic dilemmas in the ICU, leverage deeply characterized cohorts, and employ a rigorous post-hoc LRTI adjudication approach incorporating multiple physicians. Importantly, clinicians with access to a HIPAA-compliant GPT-4 interface can readily use our prompt without any prior bioinformatics expertise. Weaknesses of this study include a relatively small sample size, assessment of a biomarker not yet commonly used in clinical practice, and restriction to mechanically ventilated patients.

Future work can test whether GPT-4 can improve the marginal performance of widely available clinical biomarkers such as C-reactive protein, assess *FABP4/*GPT-4 classifier performance in larger independent cohorts, and evaluate these methods for the diagnosis of other critical illness syndromes such as sepsis.

## Author Contributions

Drs. N. Spottiswoode and C. R. Langelier had full access to all of the data in the study and take responsibility for the integrity of the data and the accuracy of the data analysis. Drs. H. V. Phan and N. Spottiswoode are co-first authors.

## Concept and design

Phan, Spottiswoode, Langelier

*Acquisition, analysis, or interpretation of data:* All authors

*Drafting of the manuscript:* Phan, Spottiswoode, Langelier

*Critical review of the manuscript for important intellectual content:* Lydon, Chu, Calfee

*Statistical analysis:* Phan, Spottiswoode, Langelier

*Administrative, technical, or material support:* UCSF

*Supervision:* Calfee, Langelier

