## Supplementary Appendix for "Integrating a host transcriptomic biomarker with a large language model for diagnosis of lower respiratory tract infection"

**Supplement**

**Table of Contents**


**Appendix 1**. Final GPT-4 prompt for diagnosis of LRTI *Page 2*

**Appendix 2**. Example of GPT-4 output for diagnosis of LRTI *Pages 3-6*

**eAppendix 1. Final GPT-4 prompt for diagnosis of LRTI.**

You are an AI clinician with expertise in critical care and/or infectious diseases. You are trying to adjudicate whether a patient has lower respiratory tract infection (LRTI).
Below, you will be given the most recent chest X-ray result (CXR) of a patient, and the clinician note for that patient prior to the most recent CXR. The note might contain information on the previous CXRs as well.
I want you to come to your own independent adjudication, regardless of what the clinical team thought. Therefore, in your adjudication, you should ignore the antibiotics treatment the patient was receiving in your analysis. You should also ignore the clinical team's LRTI diagnosis in your analysis.
I want you to give an answer in two sections. In the first section, called "Analysis", you analyze the provided information step by step to determine if the patient has LRTI, and present your analysis in a numbered list format. In the second section, called "Adjudication", you weigh the evidence for and against LRTI, then decide the patient is more likely to have LRTI or not by choosing one of two options, "LRTI" or "no LRTI", to be your final conclusion.

Here is the CXR:
*[copy paste the CXR from the docx file here, remove any trailing new lines at the end of the text]*

Here is the note:
*[copy paste the note from the docx file here, remove any trailing new lines at the end of the text]*

**eAppendix 2. Example of GPT-4 output for diagnosis of LRTI.**

**Patient 409**

| |
|---|
| **Versa Chat settings:**<br>Engine: Azure OpenAI<br>Data Source: OpenAI Web<br>Model: GPT-4 Turbo<br>Temperature: 0.2<br>Conversation mode: enabled |

<u>Attempt 1</u>

Analysis:

1. **Chest X-ray (CXR) Findings:**
   - The most recent CXR indicates the presence of a left lower lobe collapse.
   - There is also a note of worsening right lower lobe consolidation, which could be due to aspiration or pneumonia.
2. **Clinical History:**
   - The patient has been admitted to the hospital for an extended period and has had multiple interventions, including intubation and extubation, valve adjustments, and surgeries related to neurocysticercosis and ventriculoperitoneal shunt (VPS) complications.
   - The patient experienced a PEA arrest, which could have led to periods of hypoxia, potentially predisposing them to infections.
   - The patient has had a recent surgical intervention (hematoma evacuation and revision of VPS) which could be a source of infection or could complicate the clinical picture.
3. **Vital Signs:**
   - The patient's temperature has remained within normal limits, which may not support the presence of an active infection.
   - The heart rate is elevated (126 bpm), which could be a sign of systemic stress, possibly due to infection.
   - Respiratory rate is within normal limits.
   - Blood pressure is slightly elevated.
   - Oxygen saturation is at 95%, which is at the lower end of the normal range and could indicate some respiratory compromise.
4. **Physical Exam:**
   - The patient is nonverbal, which limits the ability to obtain symptoms directly related to respiratory distress or infection.
   - Neurological status seems to be the focus of the patient's assessment and care, given the history of neurocysticercosis and recent neurosurgical interventions.
5. **Laboratory and Other Clinical Information:**
   - There is no mention of white blood cell count or other laboratory markers that could indicate infection.
   - The patient is on dexamethasone, which could mask signs of infection due to its anti-inflammatory effects.
   - There is no mention of sputum culture or other microbiological tests that could confirm the presence of an LRTI.
6. **Other Considerations:**
   - The patient's history of neurocysticercosis and recent neurosurgical interventions could lead to an immunocompromised state, increasing the risk for infections, including LRTI.

- The presence of an endotracheal tube and recent surgeries could increase the risk of nosocomial infections.

Adjudication:
Based on the analysis above, the evidence suggesting the presence of an LRTI includes the worsening right lower lobe consolidation noted on the CXR, which could represent aspiration or pneumonia, and the elevated heart rate which might indicate systemic stress from an infection. However, the lack of fever, normal respiratory rate, and the absence of leukocytosis or purulent sputum in the information provided make the diagnosis of LRTI less certain.
Considering the balance of the evidence, the patient is more likely to have an LRTI, primarily due to the radiological findings of consolidation and collapse, which are consistent with this diagnosis, despite the absence of some clinical signs typically associated with infection.
Therefore, the final conclusion is:
**LRTI**

<u>Attempt 2</u>
Analysis

1. **Chest X-ray (CXR) Findings:**
    - The CXR from **[date redacted]** shows an endotracheal tube in place, terminating 34 mm above the carina, which is appropriate placement.
    - There is an interval development of left lower lobe collapse since the comparison CXR on the same day.
    - There is worsening right lower lobe consolidation, which could be due to aspiration or pneumonia.
    - The cardiomediastinal silhouette remains unchanged.
2. **Clinical History:**
    - The patient was admitted on **[date redacted]** and has had a complicated hospital course including intubation, extubation, ventriculoperitoneal shunt (VPS) replacement, a PEA arrest with successful resuscitation, and multiple surgical interventions for neurocysticercosis and its complications.
    - The patient has had a recent surgery for hematoma evacuation and revision of the VPS on **[date redacted]**
3. **Vital Signs:**
    - The patient's temperature has been within normal limits.
    - The heart rate is elevated at 126 bpm.
    - Respiratory rate is within normal limits at 18 breaths per minute.
    - Blood pressure is at the higher end of the recorded range but not hypertensive.
    - Oxygen saturation is at 95%, which is within normal limits but on the lower end of the expected range for someone on supplemental oxygen.
4. **Physical Exam:**
    - The patient is nonverbal, which could be due to underlying neurological conditions or sedation.
    - Pupils are sluggishly reactive, which could indicate central nervous system involvement.
    - The patient is following commands with the right upper extremity and moving both lower extremity toes spontaneously.
5. **Problem-based Assessment & Plan:**
    - The patient has a history of neurocysticercosis with recent neurosurgical interventions.
    - There is mention of cerebral edema being treated with dexamethasone.
    - The head of the bed is maintained at >45 degrees, likely for both neuroprotective reasons and aspiration prevention.

- Tube feeds have been restarted, indicating the patient is receiving enteral nutrition.

6. **Other Considerations:**
   - The patient has had a recent PEA arrest secondary to a pulmonary embolism, which could predispose to aspiration events.
   - The patient is on dexamethasone, which can suppress immune responses and increase the risk of infection.
   - The patient's recent tachycardia and hypertension were relieved by a bowel movement, suggesting these symptoms were not due to infection.

Adjudication

Based on the analysis of the provided CXR and clinical notes, the evidence for LRTI includes the worsening right lower lobe consolidation and the new left lower lobe collapse. The patient's elevated heart rate could also support the presence of an infection. However, the normal temperature and the fact that the patient's tachycardia was relieved by a non-infectious intervention suggest that the heart rate may not be due to an infection. The unchanged cardiomediastinal silhouette does not provide additional support for LRTI.

Given the evidence, the patient is more likely to have **LRTI** due to the radiographic findings of worsening consolidation and new collapse, which are consistent with infectious processes in the lung, especially in the context of recent aspiration risks and immunosuppression from steroids.

<u>Attempt 3</u>

Analysis

1. **Chest X-ray (CXR) Findings:**
   - The CXR indicates that there is a new development of left lower lobe collapse.
   - There is also a noted worsening of right lower lobe consolidation, which could be due to aspiration or pneumonia.
2. **Clinical History:**
   - The patient has a complicated medical history with neurocysticercosis, multiple shunt failures, and a pulmonary embolism (PE) arrest.
   - The patient has undergone multiple surgical interventions, including ventriculoperitoneal shunt (VPS) revisions and hematoma evacuation.
   - The patient has been intubated and extubated multiple times, which increases the risk for aspiration and subsequent pneumonia.
3. **Vital Signs:**
   - The patient's temperature is within normal limits, which may or may not rule out infection.
   - Tachycardia is present (heart rate of 126), which can be a sign of infection but can also be due to other causes.
   - The respiratory rate is within normal limits.
   - Blood pressure is slightly elevated, which could be due to various reasons, including stress or pain.
   - Oxygen saturation is at 95%, which is at the lower end of the normal range and could suggest some degree of respiratory compromise.
4. **Physical Exam:**
   - The patient is nonverbal, which limits the ability to obtain symptoms such as cough or dyspnea.
   - Neurological status is compromised due to underlying neurocysticercosis and recent surgeries, which may mask typical signs and symptoms of LRTI.
5. **Recent Interventions and Treatments:**

    - The patient has had a recent external ventricular drain (EVD) placement and revisions of the VPS, which can be associated with infection risk, but not specifically LRTI.
    - The patient is on dexamethasone, which can suppress the immune response and potentially mask or exacerbate an infection.
    - The patient has had recent mobilization and physical therapy, which can help prevent respiratory infections by improving lung expansion and secretion clearance.
6. **Laboratory and Imaging:**
    - No recent blood work is provided to assess for markers of infection such as white blood cell count or inflammatory markers.
    - The most recent head CT and MRI do not provide information about the lower respiratory tract.
7. **Risk Factors for LRTI:**
    - The patient has several risk factors for LRTI, including recent intubation, neurosurgical procedures, and a history of PE arrest, which could predispose to aspiration.

Adjudication

Given the evidence presented:

- The new development of left lower lobe collapse and worsening right lower lobe consolidation on CXR are radiographic findings that can be associated with LRTI.
- The patient's history of intubation and neurosurgical procedures increases the risk for aspiration pneumonia.
- The absence of fever does not rule out LRTI, especially in the context of immunosuppression due to steroids.
- Tachycardia could be a nonspecific sign of infection.

**Conclusion: LRTI**

The radiographic findings, combined with the patient's clinical history and risk factors, support the diagnosis of a lower respiratory tract infection more than any alternative diagnosis provided in the information.
